# ‘It’s a bit of a mess’: the impact of later alcohol trading hours for bars and nightclubs in Scotland according to qualitative interviews with local stakeholders

**DOI:** 10.1101/2025.10.31.25339073

**Authors:** Gemma Mitchell, Karen J Maxwell, Rachel O’Donnell, Megan Cook, Isabelle Uny, Carol Emslie, James Nicholls, Niamh Fitzgerald

## Abstract

**Background and aims:** Permitting alcohol outlets to trade later at night is associated with increased intoxication, violence and burden on public services. Between 2017 and 2019 two processes led to later trading hours in 10 nightclubs in Glasgow (1-hour pilot extension, to 4am) and 38 bars in Aberdeen (1-3 hours extension, to 3am). We explored stakeholder views on the impact of these later hours against the backdrop of the Covid-19 pandemic and UK cost of living crisis.

**Methods:** Semi-structured interviews conducted March 2022 to March 2023 with 39 stakeholders (10 public health/community/police; 14 frontline services; 9 licensing stakeholders (non-trade); and 6 venue owners/managers), analysed using framework analysis.

**Results:** Participants across both cities reported mixed consumer demand for extended trading hours. Consequently, later hours were used irregularly and unpredictably by venues and economic benefits were at best smaller than anticipated. Low demand was attributed to the pandemic, cost of living crisis and changing drinking patterns. Frontline workers were more likely than those in other/more senior roles in policing, health and local government to report increased violence and disorder during the later hours. In Aberdeen, changes were perceived broadly negatively, due to greater demand from often vulnerable drinkers later at night, which outstripped the capacity of ambulance, policing, transport and voluntary services. In Glasgow, impact on services was more often described as mixed or neutral, although some transport and voluntary services were not available to cover the extra hour.

**Conclusions:** Night-time economy stakeholders in Glasgow and Aberdeen perceived little or no economic benefits from the introduction of later trading hours, lower than anticipated consumer demand and increased demand for services, which often outstripped service capacity. Policymakers should treat with scepticism claims of benefits from later trading hours and consider other measures which address alcohol harms and pressure on services.

**KEY POINT SUMMARY:**

- Demand for later hours was mixed in both cities; subsequently, use of the extensions by venues was unpredictable and reported economic benefits minimal.
- The after-effects of the Covid-19 pandemic, cost of living crisis and changing drinking patterns were described as factors influencing lower than expected demand.
- Frontline workers were more likely than those in strategic roles to describe increased violence and disorder during the later hours.
- Ambulance, policing, transport and voluntary services struggled to meet demand from often vulnerable drinkers during the later hours in Aberdeen, and some transport and voluntary organisations were unable to meet demand during the extra hour in Glasgow.

## INTRODUCTION

Effective approaches to addressing alcohol harms include reducing the affordability, marketing and availability of alcohol (1), although the mechanisms underpinning, and scale of, the effects of availability on harms are unclear (2–4). Availability includes temporal (trading hours of outlets) and spatial (outlet density) availability (5); it is the former that is the focus here. There is a fairly consistent link between later trading hours (particularly for on-trade premises, i.e. bars, pubs and clubs) and increased alcohol consumption, injuries, assault and burdens on public services (6–11). Conversely, a reduction in hours for on-trade premises is associated with a decrease in harms (11– 14) that sustains over time (15). Many studies, however, do not clearly distinguish between premise type, there is a tendency to focus on acute rather than long-term harms, and there are few studies which give in-depth insights into the experiences of staff on the ground when trading hours change (16).

In the UK, alcohol availability is regulated through three systems of premises licensing which differ significantly (17,18). In Scotland, under the 2005 Licensing Act (‘the Act’) and subsequent legislation, local Licensing Boards are the decision-making body on hours of sale for on-trade licensed premises (where alcohol is consumed in the venue selling it), although there is a strong statutory presumption against 24-hour licences. These Boards publish licensing policy statements setting out their standard licensing hours (in some cases for premises of different types). Trading hours for new premises and requests for additional hours for existing premises are considered by the Licensing Board and are generally granted unless deemed likely to be detrimental to one of five statutory licensing objectives. In Scotland these objectives are: preventing crime and disorder, securing public safety, preventing public nuisance, protecting children and young people from harm and protecting and improving public health (18). Statutory consultees (and members of the public) can make representations or objections to inform licensing decision making; these include the police, local health authorities, fire authorities, and community councils.

Changes in trading hours as a result of the 2005 Act have not previously been evaluated in Scotland and the association between temporal availability and harms in the UK has not been entirely clear (16). The Licensing Act (2003) in England/Wales introduced a presumption in favour of granting late-night extensions (including up to 24-hour opening) unless objectors could argue that to do so would contravene specific licensing objectives. The most robust evaluation of this change found that later trading hours did not lead to increased rates of violence in Manchester, although the patterning of weekend violence changed such that there were more incidents later at night (36% increase between 03:00 and 06:00) (19). Further, though no change in overall consumption was found, drinking occasions shifted to slightly later at night and involved preloading (where alcohol is consumed first in off-trade then on-trade) compared to Scotland after the introduction of the Act (20). Other studies of the 2003 changes were generally of weaker design or quality (21). A few qualitative studies focused on the perceptions of stakeholders in strategic roles (e.g. in licensing authorities or police services), who generally felt that alcohol-related problems had remained stable or declined following the introduction of the Act (22).

Between 2017 and 2019, two different processes (23) led to later opening hours for 10 nightclubs in Glasgow (a 1-hour pilot extension) and 38 bars in Aberdeen (1-3 hours extension, up to 3am). These policy changes took place at a time of economic and social instability in the UK. Pre-2020, health and social care systems had been under strain following over a decade of austerity (24,25). From 2020, the Covid-19 pandemic compounded existing public health crises (26) and from late 2021, the UK experienced a cost-of-living crisis due to rising inflation and a fall in real disposable incomes (24). Given that these contextual factors are likely to have independent and interdependent impacts on the outcomes and services affected by late night alcohol sales, we aimed to explore stakeholder views on the impact of the extended hours against the backdrop of the Covid-19 pandemic and UK cost of living crisis.

## METHODS

### Overview

We present findings from 39 semi-structured interviews with local night-time economy stakeholders in Glasgow and Aberdeen, which comprised part of the ‘Evaluating Later or Expanded Premises Hours for Alcohol in the Night-Time economy (ELEPHANT) study.

### Sampling and recruitment

The sampling frame was developed via discussions within the study team and local stakeholders in public sector licensing roles, then via snowball sampling. We aimed to recruit 60 participants from the following groups: public health and community planning, stakeholders dealing with the public in frontline roles (e.g. police, NHS Accident and Emergency staff, third sector workers), non-trade licensing stakeholders (e.g. Licensing Board members, licensing police, licensing lawyers) and venue owners/managers. Within those groups, we sampled for variety related to individual role, experience, and seniority; and for venues’ location, type, clientele, and trading hours. Of the hundred and twenty-two individuals approached, 46 agreed to participate and 39 participated in interviews (four were uncontactable to arrange interviews and three did not attend scheduled interviews). Recruitment proved challenging, particularly amongst the venue owners/ managers group, where, for example, we were only able to recruit four out of 33 individuals approached in Aberdeen. We ended recruitment in agreement with the study steering committee after pursuing all available recruitment leads. A further study focused solely on trade views of later extensions to bar opening hours in Glasgow reports more in-depth findings from that perspective (27).

### Data collection

Thirty-six interviews were conducted with 39 participants by telephone or online (by RO, IU, KM, GM, MC and NF; three were paired interviews) between March 2022 and March 2023 (see Table 1). Separate topic guides were developed for each stakeholder group, each covering: views on rationale for the changes, impact of the later hours, impact of COVID-19 restrictions, and the future of the night-time economy. Each topic guide was based on alcohol licensing literature and team discussion. Interviews lasted between 38 and 90 minutes (median = 66) and were audio-recorded and transcribed by a transcription agency.

**Table 1:**
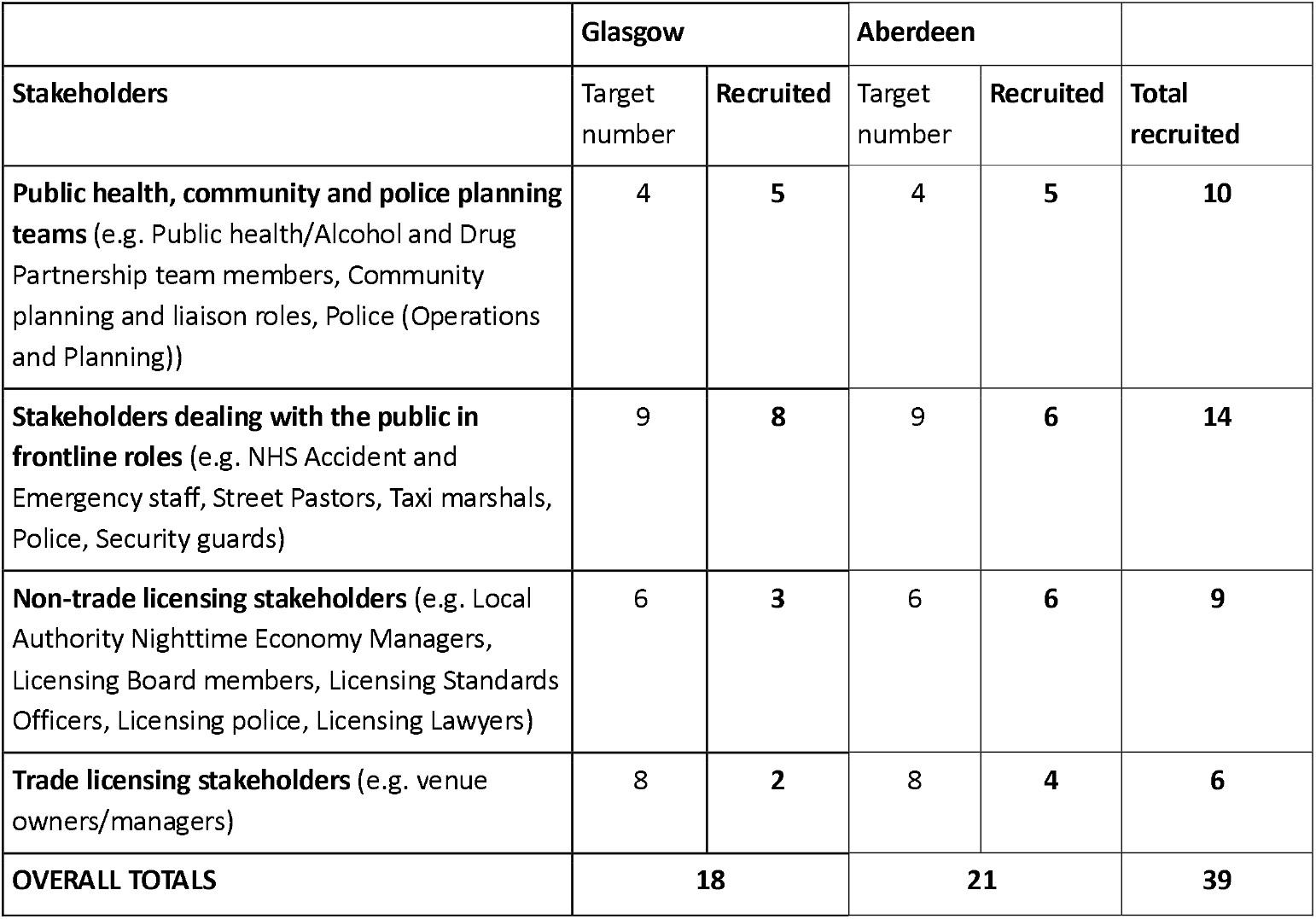
Characteristics of interview participants.

### Analysis

Data were analysed (by GM, RO) using the framework approach (28,29). This approach is well-suited to the analysis of in-depth interview data, as summarisation of data into a framework grid within the NVivo programme facilitates analysis across cases without losing the wider context of each participant’s account (28). First, the interviews were de-identified, which helped with familiarisation (GM). A thematic framework was developed (by RO and GM), based on deductive (considering the topic guide) and inductive (reading transcripts) techniques. Data summaries were written in relevant cells of the framework grid (GM and RO), incorporating hyperlinks to transcripts to facilitate data retrieval. These were used to identify high-level themes (GM, RO, MC) before further in-depth analysis was conducted (GM). We focused on stakeholder experiences of the changes and the impact of the pandemic and cost of living crisis, with the former deriving from deductive and the latter derived from inductive techniques. GM, RO and MC refined themes based on re-examining data and reflexive team discussions.

### Ethics

Informed consent was provided by all participants and any identifying information removed. The ELEPHANT study was approved by the University of Stirling Ethics Committee for NHS, Invasive or Clinical Research (NICR 0457).

## RESULTS

Participants across both cities reported that any economic benefits of extended hours were at best smaller than anticipated, as consumer demand was mixed. Lower than anticipated demand was attributed to the pandemic, UK cost of living crisis and long-term changes to drinking patterns. We report on the impacts on businesses, customers and public services.

### ‘It’s a bit of a mess’: mixed demand and minimal economic benefits for businesses

Most participants, including several trade representatives, reported that consumer demand to attend venues later at night was mixed across both cities, and the economic benefits of the changes minimal. The mixed demand for the later hours was surprising for many participants, including some venue representatives:

> *We assumed we’re going to be busy. Mobbed. ‘Til four am. But what happens is some of these nights, [the venue] wasn’t mobbed. (Participant 29, licensing (trade), Glasgow)*

Changes to people’s drinking patterns, particularly among young people and students, were reported to have led to this lower than expected demand in both cities. In Glasgow, several participants suggested that fewer people were travelling to the city centre post-pandemic, because they had lost the habit and had less disposable income due to the cost-of-living crisis, or because young adults had never been ‘used to it’:

> *I think it’s just culture. ‘Cause they’ve had years of not getting out [during the pandemic], a lot of folk just won’t do it now. A lot of folk that have turned eighteen, nineteen, twenty have never been in a club. So they’re not used to it. (Participant 14, police (frontline), Glasgow)*

> *I think COVID has absolutely thrown a bomb right into the middle of it. It has changed why people come out to night-time economy. I think part of it is because of what’s happening in terms of, if you come into the city, can you get home? […] Then on top of that you’ve now got, can you afford it? (Participant 38, public health and community planning, Glasgow)*

In Aberdeen, participants added that the local economy had not recovered from the pandemic, and was struggling pre-pandemic, in part due to the struggling gas and oil sector. These factors combined with other changes to reduce demand:

> *I got a sense that the drinking culture was changing in any case, in terms of, Friday nights became very quiet. And whether this was some of the oil downturn which obviously was really impactful on Aberdeen. Or the cost of alcohol. I think the social media generation changed things a little bit as well, in terms of a lot of socialising was done behind closed doors. So the preloading was really coming on board [at the time of the changes]. (Participant 30, police (strategic/planning), Aberdeen)*

A combination of lower than anticipated demand and additional costs of keeping venues open til later at night influenced perceptions of whether there were economic benefits from later trading hours. Venues reported that the extra trading hours helped them ‘tread water’ and/or provided minimal, if any, economic benefits to them:

> *There is a cost [associated with the later hour]. Your set up, your lights are still on and your payroll…So giving yourself more scope doesn’t give yourself more profit… It [lack of profit] doesn’t mean drop it [the later hour], it means [a need to consider] sustainability [of later trading]. (Participant 29, licensing (trade), Glasgow)*

> *The economic benefits [are] very marginal… [in terms of] how much people spend on any given night. Arguably, it’s probably not worthwhile, in fact, it might be even less worthwhile than closing earlier. (Participant 28, licensing (trade), Glasgow)*

There were mixed reports of whether there were extra costs specifically associated with later trading, over and above the standard costs of trading (staffing, lighting etc.). Glasgow-based venue representatives and non-trade licensing stakeholders reported that extra licensing conditions were placed on all nightclubs taking up the extra hour which created extra costs (for example, door stewards and additional first aid resources). Aberdeen-based venues which received extensions to trading hours reported few equivalent costs as additional licensing conditions were not routinely applied.

In Aberdeen, increased competition was noted because pubs and bars could open later without necessarily incurring the same costs as nightclubs. Aberdeen-based participants reported hybrid and nightclub venues removing entrance fees to remain competitive, with associated loss of revenue for some venues:

> *It’s really bad from the night-time economy point of view; it has a massive effect on club-run venues. Because now that bars are open [later] and any bar can essentially be a club, it makes running a club a lot harder – it makes people not willing to pay things like an entry charge. (Participant 25, licensing (trade), Aberdeen)*

Additionally, where nightclubs in both cities did charge a fee, venue representatives reported having to stay open regardless of how busy they were because customers had paid to stay until a later time, which incurred costs and could be unprofitable.

One venue representative in Aberdeen reported clear economic benefits for their venue, stating that the later hours created extra revenue that had helped them stay open in tough economic conditions. One Glasgow nightclub representative reported that although the direct economic benefits were minimal, the 4am closing time kept the venue competitive internationally, helping to recruit music artists:

> *The key benefit for a premises such as ours where we bring artists from all around the world to play in our premises…these are people that are used to maybe playing in other cities for ten hours solid and they were coming to Glasgow and they were only being able to play for two hours. So [the changes] opened up [the ability to recruit leading artists]. (Participant 28, licensing (trade), Glasgow)*

Yet, the combination of lower demand and costs meant that the majority of venue representatives across both cities, as well as many other participants, reported that venues had to weigh up the financial viability of staying open later, and at times, deemed it unviable. Consequently, the trading hours and usage of extended hours of any given premises in either city on any given night, was unpredictable and ‘messy’:

> *It’s a limited number that use the four o’clock [extension]…it’s not beneficial to them…they’re not getting the numbers through the door anymore. (Participant 14, police (frontline), Glasgow)*

> *So the question was ‘has it [extended hours] worked?’ I think it’s a bit of a mess, to be honest. (Participant 27, licensing (trade), Aberdeen)*

### ‘There’s quite a lot of police presence’: impacts on violence, disorder and behaviour

In Aberdeen, most participants thought the changes had a negative or mixed impact on alcohol harms, including increased violence and disorder during the later hours. Most participants in Glasgow had mixed views or thought the changes had little impact on levels of violence and disorder, which was partly attributed to the small size of the pilot in the city (only affecting 10 venues). For frontline service providers in both cities (and some planning representatives), the last hour of venue opening was noted as the worst time for harm, with each hour later perceived as leading to higher levels of violence and disorder:

> *Whereas before, [violence and disorder] might have been a little bit earlier, it’s now later…We are finding quite a lot of incidents in the evenings. More of the same incidents are happening on a night-to-night basis, rather than one or two isolated incidents… There’s quite a lot of police presence, and maybe there wasn’t so much of that back then. And I feel it’s needed, to be fair. (Participant 23, public health, community planning, Aberdeen)*

> *We certainly noticed that we were still getting the more extreme situations with people right through [the night]. Whereas [previously] there would be a lull after three o’clock, that lull doesn’t happen anymore. (Participant 2, frontline service provider, Glasgow)*

Both the above participants (and others) linked this to the after-effects of the pandemic, with increased levels of intoxication by younger night-time economy users after pandemic-related restrictions ended.

Many participants across all categories noted that meeting the needs of vulnerable drinkers during the extra hours posed specific challenges, because demand for services continued after venues closed:

> *Our resource is usually pretty stretched at that time [after midnight]. Because what happens a lot of the time is, night-time economy [users] come out, from midnight ‘till three a*.*m*., *and they go back to their communities. And then quite often, we get calls to house parties, or disturbances, and house domestics, or whatever. So, there is that spill over. (Participant 30, police (strategic/planning), Aberdeen)*

### ‘The police are being used as a late-night taxi service’: public resources stretched thin and at times unable to provide services during later hours

Despite lower than anticipated demand for the later hours, many participants in all categories across both cities reported that frontline service providers such as police, ambulance, transport and voluntary services struggled to meet the needs of vulnerable drinkers during the additional hours. The impacts of this were perceived differently in each city, with Aberdeen-based participants more likely to say that the impacts on services were significant and due to the changes in trading hours:

> *[The later hours cause] a huge demand, strain, I will use the word strain, ‘cause it is, particularly over the weekends for staff. (Participant 12, public health and community planning, Aberdeen)*

Several participants, mostly those based in Aberdeen, raised concerns about frontline workers having to cover roles usually performed by other services at the later hours, thus changing their core responsibilities:

> *With later opening venues, and particularly, the longer [at night] premises open later, the more there is an expectation and a reliance on our emergency services…that impacts, then, on the NHS… [If] you’re incapacitated in the city centre, unfortunately, it tends to fall onto the police…we are dealing with medical matters, because the ambulance service is not [able to respond]. (Participant 20, police (frontline), Aberdeen)*

> *That extra hour between two and three, I’ve had licensees say to me that, for that extra hour, I’m basically just a social worker and I’m organising taxis…my job has become not one of sell[ing], it’s dealing with the fall out. (Participant 35, licensing (non-trade), Aberdeen)*

Shifting priorities after the pandemic and increased violence during the later hours led to fewer people willing to work as taxi drivers or marshals at these hours, according to several participants. This combination of limited resources and demand from vulnerable drinkers during the later hours led to a widespread expectation that the shifts of police officers and paramedics would finish several hours after venue closing times, particularly in Aberdeen. Further, frontline service providers reported that demand in city centres led to fewer staff and resources being allocated to other locations, rather than additional resources being provided to service the extra trading hours. The extra demand on services lasted well beyond the actual closing time of premises and into subsequent days, according to many participants:

> *We see a lot of our patients on a Sunday who present throughout the day, and it’ll be a result of something that happened on Saturday night, or on Friday. So, they may well have been associated with clubs, and pubs*… *but the impact of that is not felt exclusively on the Saturday night, Sunday morning, it’s there for the next twenty-four hours as well. (Participant 5, NHS Accident and Emergency doctor (frontline), Aberdeen)*

For participants based in Glasgow, the impacts of the later hours on already stretched services were viewed by many as minimal because of lower than anticipated demand and the limited nature of the pilot in only ten venues, which were considered good operators:

> *The [venues] that got the late[r] licence were already dealing with everything correctly anyway so that there wasn’t a big impact because there wasn’t a lot of issues in their clubs. But that’s them, you don’t know what the rest of [venues] were like because [there are many] nightclubs in the city centre and [the extended hour] was only given out to a handful of them. (Participant 15, police (frontline), Glasgow)*

Despite many Glasgow-based participants describing the impact of the later hours on services as minimal, services such as bus and rail, voluntary organisations and taxi marshals were unable to provide any services during the extra hour between 3 and 4am. A transport representative reported they were not involved in the consultation on the later hours pilot, but were already limited in their ability to provide a service and extra transport provision in the later hours was not commercially viable. Similarly, train services did not operate during the extended hours in the city, with existing services experienced as affected by staff shortages and strikes. This was reported to have led to increased demand for taxis, whilst taxi availability was already reduced due to the pandemic. The lack of transport was seen as suppressing consumer demand. Aberdeen-based participants also reported a lack of transport during the later hours, with a lack of taxi provision in both cities also described as putting pressure on frontline services, leading workers to, again, perform roles outside their core responsibilities:

> *It’s almost like the ambulance is being used, or I would say, the police are being used as a late-night taxi service because there’s no other mode of transport. (Participant 25, licensing (trade), Aberdeen)*

Each later hour that premises remained open at night was reported to make a difference to whether services could be provided, especially in the time customers would be on the streets after premises closed. In Aberdeen, some transport providers and voluntary services were able to cover the extra bar hours, partly because nightclub closing hours had always been 3am in the city. In Glasgow and Aberdeen, taxi marshals are contracted by city councils to manage taxi ranks – ideally until an hour after premises closed. Yet, in Glasgow, a city council representative reported that the council could not extend taxi marshal provision until 5am because it was unaffordable. Again, in Glasgow, the extra hour after clubs closed at 4am was reported by a voluntary frontline service provider as impossible to cover:

> *Clearly if clubs are staying open until four o’clock, stopping [our service provision] at four o’clock would seem not the right thing to do. But equally we needed to look at how our organisation could then respond to that? Because it’s quite one thing working till four o’clock in the morning, which essentially it isn’t four o’clock because [the work doesn’t stop at exactly 4am]. But it’s quite another thing to say five o’clock in the morning. (Participant 2, frontline service provider, Glasgow)*

The third sector organisation this frontline service provider represented ultimately made the decision that they could not provide services post-4am, as it was not sustainable.

Uncertainty about shift patterns due to unpredictable usage of extended hours was also reported by several Aberdeen-based frontline service providers, with the impact described as affecting body clocks and family time, alongside the increased exposure to violence and disorder:

> *I do remember our shifts having to get adjusted ‘cause we used to work the late shift [until 1am]… and initially that was dealt with by offering overtime, which is a cost to the public purse, and the negative impact on that is that you’re not getting home until five o’clock in the morning on a Saturday morning or a Sunday morning. And those with family, that has a social impact… and work/life balance gets impacted as well. (Participant 17, police (frontline), Aberdeen)*

There was a disconnect between the views of frontline staff expressed above, who reported negative impacts of the changes, and those in strategic roles in public services or non-trade licensing roles (including those responsible for implementing local licensing policies). The latter reported mixed, neutral or minimal impacts on alcohol-related harms, and/or viewed the later hours as positive in terms of harm reduction and boosting their city’s night-time economy. Some explained their perception of minimal impact as being due to the lack of use of the extra hours and lower demand.

> *The closing of bars and nightclubs at midnight, two in the morning, three in the morning, definitely benefited us. Because taxi ranks didn’t have the same influx, you know, that exodus at the same time. (Participant 30, police (strategic/planning), Aberdeen)*

> *Initially it was a bit of a concern. But actually, when it happened, and the things that we were concerned about didn’t happen, I think we almost forget that the clubs were opened that hour. And I think, also from speaking to some of the venues, they didn’t stay fully open [didn’t always use the extra hour]. (Participant 8, public health and community planning, Glasgow)*

## DISCUSSION

### Summary of findings

Participants across Glasgow and Aberdeen reported that consumer demand to stay in premises later at night was lower than expected; consequently, venue use of granted extensions was unpredictable and economic benefits reported as minimal in both cities. Lower than expected demand was linked to the after-effects of the pandemic, the cost-of-living crisis, lack of public transport late at night, and changing drinking patterns. For Aberdeen-based participants, the changes were perceived by most as having negative impacts, stretching frontline service capacity, but services were reported as able to cover the extra hours. In Glasgow, most participants reported mixed or neutral impacts, although one frontline voluntary sector service and most transport providers were noted as unable to cover the later hour. Frontline service providers were more likely than those in planning/strategic roles to report concerns about impacts on the safety and wellbeing of both night-time economy customers and workers.

### Comparison with prior literature

Few peer-reviewed studies have specifically focused on falling demand for late-night alcohol venues, but the trend has been widely reported in contemporary media (30,31). The explanations given by our participants accord with evidence of falling real disposable income (32), reduced availability of public transport (33) and reduced alcohol consumption especially amongst young people (34,35). Whilst preloading (drinking alcohol in domestic settings prior to attending on-trade premises) has been increasing generally (36), it is not clear that this would have actually reduced spending in premises (as our participants believed). Pre-drinkers in Canada consumed more alcohol in on-trade premises than non-pre-drinkers (37), whilst other studies find that pre-drinkers tend to be more intoxicated and at higher risk of harm (38–40).

In Queensland, Australia, restrictions to late night alcohol sales (service stopping at 3am instead of 5am), were reported by some bar owners as having caused their businesses to lose money (41). Many others echoed our findings that they rarely used, or did not use, their later licences anyway prior to the changes, were quiet during the later hours, or that closing earlier saved staff costs or had minimal negative impact. In the same qualitative study, police and voluntary services reported a range of benefits to earlier closing including reduced alcohol-related anti-social behaviour, drunkenness and injury, as well as a general sense of a safer, more positive environment around late-night venues (41). In a separate observational study of bars in five Australian cities, customer numbers fell later at night, whilst the intoxication level of remaining customers continued to increase (42).

Our findings give voice to frontline workers’ experiences that echo previous quantitative studies finding negative impacts of later alcohol trading hours on alcohol harms including violence (8,9)and ambulance call-outs (43,44). In qualitative interviews about the impact of alcohol on the Scottish ambulance service, paramedics felt that alcohol-related calls placed significant pressure on the service (45). They also reported relatively routine experiences of aggression and harassment when dealing with calls linked to alcohol and in late-night premises, that did not emerge as a theme from frontline services staff in this study – perhaps because we did not specifically ask about individual experiences. However, participants, particularly in Aberdeen, were concerned about the impact of later trading hours on the safety and wellbeing both of consumers and hospitality staff, both of whom risked exposure to violence, and struggled to get transport home. These findings echo local reports (46), as well as survey findings and other studies summarised by Alcohol Action Ireland (47), yet the health and safety of frontline service or hospitality workers were not considered when the changes were first proposed (23).

Participants in Aberdeen tended to report greater negative impacts of the later hours than those in Glasgow. These perceptions mirror our quantitative findings (see (48)) that found a significant increase in weekend night-time alcohol-related ambulance attendances, and in recorded crimes in Aberdeen, and no significant impacts on these two outcomes in Glasgow. Our interpretation of these findings together is twofold: firstly, that the number of premises (n=10) granted one extra hour in Glasgow was small relative both to the premises numbers and degree of changes in Aberdeen, and to the overall number of premises in each city (with Glasgow being much larger). Secondly, our qualitative findings allow identification of impacts beyond those recorded in routine ambulance call-out and crime statistics analysed by Sheikh et al. (48): demand for ambulances which does not result in an ambulance attendance - sometimes because other services end up responding instead; police and other services providing support for vulnerable people unrelated to any crime; and lack of services after 4am leaving people to manage without support (again leaving no statistical record). Our findings therefore provide an important counterpoint to any sense that the Glasgow changes had no negative impacts on services.

### Implications for policy and practice

Most importantly, these findings illustrate that extending opening hours of licensed premises late at night is neither a harmless nor particularly effective route to generating economic or other benefits. This contradicts recent policy rhetoric in both Ireland (49) and England and Wales (50,51) which seems to be naïve about the diverse harms of continued extensions to late-night opening, and overly optimistic about the potential of such changes for driving recovery in the hospitality sector or contributing to wider economic growth. If reviving local night-time economies or high streets is a priority, governments should look beyond alcohol-focused hospitality, whilst also considering ‘sweetspot’ pricing and online availability policies, likely to reduce harm whilst having minimal negative impacts on businesses (52).

Furthermore, the findings in this paper and our separate quantitative study (Sheikh et al., n.d). suggest that the scale of changes in alcohol trading hours may matter in ways not acknowledged in the current licensing system. If the degree of harms arising from later trading can be mitigated to some extent by allowing fewer venues to have those extra hours, then licensing laws would need to change to allow local areas to apply limits on the numbers of late-night venues. This would increase the economic viability of those awarded later licences, whilst minimising the impact on services.

Finally, where later opening hours are being considered despite the harms that are likely to occur, our findings point to the need to consider, and plan mitigation for, such harms. These include providing for frontline service provider and hospitality worker safety and wellbeing and resources for local late-night services including voluntary sector and transport providers. If there are any economic benefits, they could at least then be considered alongside questions of who pays to address the harms that accompany them. Otherwise, it is the public and third sector that pay the bill, whilst the potential economic benefits largely accrue to businesses.

### Implications for research

Our findings highlight the limitations of research on later trading hours that relies only on secondary data analysis. Participants reported that many of the services they provided, to vulnerable or intoxicated individuals, did not appear in any datasets such as police or health service records. Further studies should therefore plan repeat observational or qualitative data collection, to capture frontline experiences. Prior to our findings being reported, the Glasgow pilot allowing nightclubs to open to 4am studied here was made permanent in late 2023, and in a new ‘pilot’, 54 bars and pubs were granted permission to extend their trading hours from midnight to 1am (52). Both provide an opportunity to conduct further research to test the hypothesis that there is a dose-response relationship between the number of premises granted extra hours and the harms that result.

### Strengths and Limitations

This study is the first in the UK and one of very few globally to have captured the voices of frontline staff who have experienced a substantial change in local late-night alcohol trading hours. Its strengths include the use of qualitative interviews to gain a depth of understanding of the experiences of later opening hours and participation of a wide range of stakeholders across two cities allowing comparison of frontline and strategic perspectives. Comparing two cities, each implementing later trading hours differently, allowed greater insights than a single site study. The limitations of this study include, but are not limited to, recruitment challenges and a relatively long period of data collection (March 2022 – March 2023). Recruitment challenges, especially amongst bar/nightclub representatives, means these views are underrepresented. A further study in Glasgow focused solely on trade views will address this limitation (27). Interviews being conducted over a year means participants’ experiences may have varied as pandemic conditions changed, however this added confidence in the themes that remained consistent across the data collection period. Economic and social contexts will always vary and interact with changes to the licensing system.

## CONCLUSION

The introduction of later trading hours for licensed premises in Glasgow and Aberdeen was perceived to have little or no economic benefits and to have increased alcohol harms and demand for services – outstripping capacity and going beyond what is recorded in routine data. Lower than anticipated consumer demand for later drinking likely explained the lack of economic benefits; however, any future upturn in demand would be expected to result in higher levels of harm. Policymakers should treat with scepticism claims of benefits from later trading hours, economic or otherwise, and should instead look to other measures to address alcohol harms and pressure on services.

## Data Availability

All data produced in the present study are available upon reasonable request to the authors

## Acknowledgements

We thank the Evaluating Later and Expanded Premises Hours for Alcohol in the Night-time Economy (ELEPHANT) Study Steering Committee and attendees at the Kettil Bruun Society annual symposium for their comments on early findings in this paper. We would like to thank Kathryn Angus for support with referencing.

## Notes

### Competing Interest Statement

The authors have declared no competing interest.

### Funding Statement

This project is funded by the National Institute for Health and Care Research (NIHR) [Public Health Research programme (129885)]. The views expressed are those of the author(s) and not necessarily those of the NIHR or the Department of Health and Social Care.

### Author Declarations

This study was approved by the University of Stirling Ethics Committee for NHS, Invasive or Clinical Research (NICR 0457).

